# COVID-19 progression is potentially driven by T cell immunopathogenesis

**DOI:** 10.1101/2020.04.28.20083089

**Authors:** Moritz Anft, Krystallenia Paniskaki, Arturo Blazquez-Navarro, Adrian Doevelaar, Felix S. Seibert, Bodo Hoelzer, Sarah Skrzypczyk, Eva Kohut, Julia Kurek, Jan Zapka, Patrizia Wehler, Sviatlana Kaliszczyk, Sharon Bajda, Constantin J. Thieme, Toralf Roch, Margarethe Justine Konik, Thorsten Brenner, Clemens Tempfer, Carsten Watzl, Sebastian Dolff, Ulf Dittmer, Timm H. Westhoff, Oliver Witzke, Ulrik Stervbo, Nina Babel

## Abstract

**Background:** The role of cellular immunity in pathogenesis of COVID-19 is unclear and conflicting data points to insufficient or pathogenic immunity as drivers of COVID-19 progression. Here we aimed to delineate the phenotype and function of the immune system in patients with moderate, severe, and critical COVID-19.

**Methods:** In this prospective study, we included 53 patients with moderate (n=21), severe (n=18), and critical (n=14) COVID-19 manifestations. Using multiparametric flow cytometry we compared quantitative, phenotypic, and functional characteristics of circulating immune cells, SARS-CoV-2 antigen-reactive T-cells, and humoral immunity.

**Results:** Deep phenotypic profiling revealed a depletion of circulating bulk CD8^+^ T-cells, CD4^+^ and CD8^+^ T-cell subsets with activated memory/effector T-cells expressing CD57^+^, HLA-DR^+^, and the key activation and migration molecule CD11a^++^ in critical COVID-19. Importantly, survival from acute respiratory distress syndrome was accompanied by a recovery of the depleted CD11^++^ T-cell subsets including T-cells expressing CD28, CD57, HLA-DR activation/effector molecules. We further observed a stronger response of S-protein specific T-cells producing inflammatory cytokines in critical COVID-19 cases. This seemingly contradictory observation is in fact confirmation of the underlying immunopathogenesis in patients with critical COVID-19.

**Conclusion:** Our findings suggest a CD11a-based immune signature as a possible prognostic marker for disease development. Our data further reveal that increased rather than decreased SARS-CoV-2 specific T cell immunity is associated with adverse outcome in COVID-19. Tissue migration of activated effectors T-cells may constitute a crucial cornerstone in the immunopathogenesis of SARS-CoV-2 associated tissue injury.

**Trial registration:** This is a prospective observational study without a trial registration number.

**Funding:** This work was supported by grants from Mercator Foundation, the BMBF e:KID (01ZX1612A), and BMBF NoChro (FKZ 13GW0338B).

**25 Word summary:** Stronger S-protein reactivity and decreased frequency of activated memory/effector T-cells expressing CD11a^++^ suggests immunopathogenesis in critical COVID-19 mediated by tissue migration of activated effector T-cells.

## Introduction

The pandemic outbreak of SARS-CoV-2 faced the human population with tremendous health, social, and economical challenges. SARS-CoV-2 can lead to acute respiratory distress syndrome (ARDS) and multi-organ failure (1). The conventional assumption on the general immunity fitness can certainly not explain the different disease manifestation. Recent data demonstrate an association of cytokine storm including high level of interleukin (IL)-6 production with severe disease (1–3) suggesting a pathological immune dysregulation. In contrast, markedly lower immune cell numbers and decreased activation levels were associated with critical COVID-19 manifestations (4, 5). Usually, a protective role of cellular immunity controlling viral infection can be assumed (6–8). However, an overwhelming immune response after viral infections leading to cell damage and organ failure was also reported (9). It is unclear whether the diminished or hyperreactive immunity is responsible for the critical COVID-19 manifestations. Our study provides a detailed characterization of non-specific and SARS-CoV-2-reactive cellular and humoral immunity in a prospective cohort of patients with different disease severity to understand their role for COVID-19 progression.

## Methods

### Study population and design

53 patients with moderate (n=21), severe (n=18), and critical (n=14) COVID-19 manifestations were recruited into the study. The severity of infection was assessed according to the guidelines of the Robert Koch Institute, Germany, Table S1. Demographic and clinical characteristics of patients are demonstrated in Tables 1 and S2.

**Table 1.** Patient characteristics

|  | Disease Severity |  |  |
| --- | --- | --- | --- |
|  | Moderate | Severe | Critical |
| Number patients | 21 (39.62%) | 18 (33.96%) | 14 (26.42%) |
| Age (NS) | 64.6 (range: 41-89) | 73.9 (range: 56-92) | 64.9 (range: 26-86) |
| Gender (Male/Female) | 10/11<br>(47.62/52.38)<sup>NS</sup> | 10/8<br>(55.6%/44.4%)<sup>NS</sup> | 13/1<br>(92.9%/7.1%)<sup>**</sup> |
| Comorbidities |  |  |  |
| Cancer | 1 | 3 | 1 |
| Chronic renal Disease | 0 | 2 | 0 |
| Obstructive Lung Disease | 2 | 1 | 0 |
| Diabetes | 1 | 3 | 3 |
| Cardiovascular disease | 8 | 6 | 2 |

Age is compared across patient categories by one-way ANOVA. Gender is compared for each patient category by $\chi^2$ . NS indicate not significant, \*\* indicate P value < 0.01

Patients with moderate and severe COVID-19 were recruited after the first symptoms occurred and a positive SARS-CoV-2 PCR result (in median 4 days after the diagnostic test) was available. For patients with critical disease, the recruitment occurred at ICU, being diagnosed with COVID-19 in median 14 days before. To ensure the comparability of the data with respect to the longer duration of COVID-19 in ICU patients, blood samples of patients with moderate and severe cases were obtained in follow up within the next 8 days after the recruitment. For 6 out of 14 critical patients, only one time point was available (Fig. 1A).

**Fig. 1:**
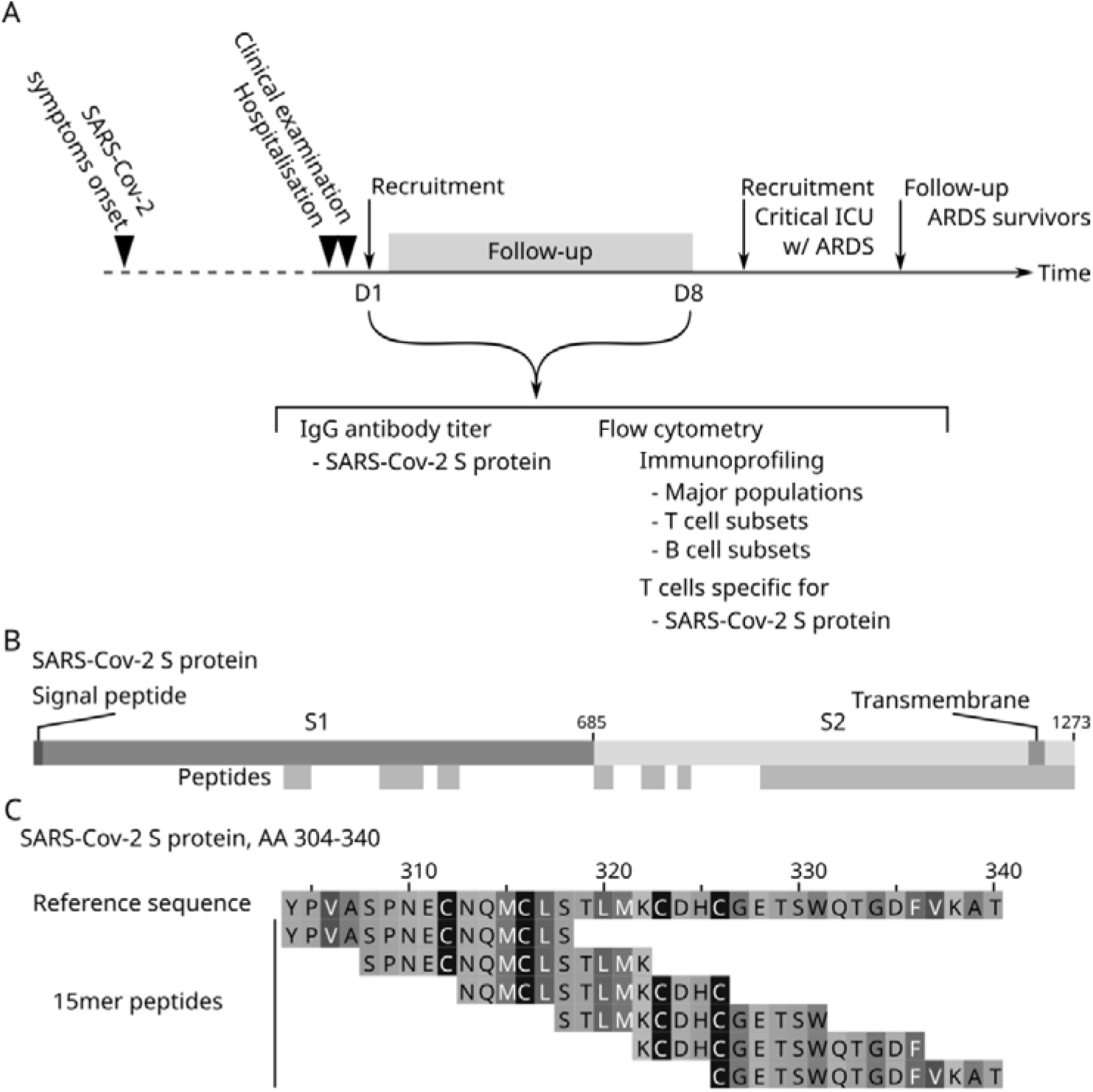
Study protocol and SARS-CoV-2 S-protein overlapping peptide pool. (A) 53 patients admitted to Marienhospital Herne – Universitätsklinikum der Ruhr-Universität Bochum, Herne, North Rhine-Westphalia, Germany and Universitätsklinikum Essen, North Rhine-Westphalia, Germany were enrolled into the study if diagnosed with COVID-19. Patients with moderate and severe COVID-19 were recruited after the first symptoms occurred and a positive SARS-CoV-2 PCR result (in median 4 days after the diagnostic test) were available. For patients with critical disease, the recruitment into the study occurred at ICU, being diagnosed with COVID-19 in median 14 days before. To ensure the comparability of the data with respect to the longer duration of COVID-19 in ICU patients, blood samples of patients with moderate and severe cases were obtained in follow up within the next 8 days after the recruitment. For 6 out of 14 critical patients, only single time point was available. The profiling included evaluation of SARS-CoV-2 S-protein specific IgG serum antibodies, as well as phenotyping of all major immune cell populations by flow cytometry, and characterization of B- and T-cell subsets. T-cells reactive to the SARS-CoV-2 S-protein were also analyzed by application of overlapping peptide pools. (B) The utilized peptide pool contained 15mer, 11 AA overlapping peptides spanning the S-protein regions 304-338, 421-475, 492-519, 683-707, 741-770, 785-802, and 885-1273 of the S-protein. (C) Principle of 15mer 11 AA overlapping peptides from SARS-CoV-2 S- protein region 304-340.

### Preparation of PBMCs and stimulation with SARS-CoV-2 overlapping peptide pools

SARS-CoV-2 PepTivator peptide pools (Miltenyi Biotec), containing overlapping peptides spanning parts of the surface glycoprotein (Fig. 1B), were used in the stimulation. The utilized overlapping peptide pool (OPP) consisted of 15mers with 11 amino acids overlap (Fig. 1C). Peripheral blood mononuclear cells (PBMCs) were prepared from whole blood by gradient centrifugation as previously described (10). Isolated PBMCs were stimulated with 1 μg/mL OPP for 16h. Brefeldin A (1μg/ml, Sigma Aldrich) was added after 2h.

### Flow cytometry

For the immune phenotyping, EDTA treated whole blood was stained as described in the Supplementary Appendix. Surface antigen of the T-cells stimulated with SARS-CoV-2 OPP were stained prior to fixation and staining of intracellular antigen, as described in Supplementary Appendix. All samples were immediately acquired on a CytoFlex flow cytometer (Beckman Coulter).

### SARS-CoV-2 IgG Antibody titers

SARS-CoV-2 IgG titers were analyzed using a commercially available ELISA (EUROIMMUN, Lübeck, Germany) per manufacturer’s instructions.

### Statistics

Flow cytometry data were analyzed using FlowJo version 10.6.2 (BD Biosciences); gating strategies are presented in Fig. S5-S9. Statistical analysis was performed using R, version 3.6.2 (11). P values were not corrected for multiple testing: Since this is an exploratory study with the aim of hypothesis generation, multiplicity adjustment is generally not recommended as relevant findings might be considered insignificant (12). In the figures, only significant P values (P < 0.05) are reported.

### Study approval

The study was approved by the ethical committee of the Ruhr-University Bochum (20-6886) and University Hospital Essen (20-9214-BO). Written informed consent was obtained from all participants.

### Results

53 hospitalized patients with moderate, severe and critical COVID-19 disease manifestations were enrolled in this prospective study. The detailed characteristics of study patients, study design and blood sampling are presented in Table 1, S2 and Fig. 1A. There were no statistically significant differences in age between the analyzed groups. Most patients in the critical groups were males. To exclude a potential bias of obtained results caused by gender mismatch between the groups, we performed a bivariate regression analysis for all significant factors. We found no evidence of a confounding effect of gender for the described markers associated with COVID-19 severity (P>0.05, Table S3).

We monitored changes of disease severity during the short follow up period. Six out of ten patients suffering ARDS dies within two weeks after study recruitment, while the remaining four ARDS patients recovered. Ten further patients showed clinical improvement and moved from the severe to the moderate disease cohort in the follow up visit.

### Degree of lymphopenia is similar across disease severity

The absolute counts of circulating leukocytes including lymphocytes, granulocytes, and monocytes were for most patients below the reference level at the first and at the follow-up visit (Fig. S1 and S2). Eosinophil counts were increased in critically ill patients (Fig. S1G). We further characterized different subsets within T- and B-cell compartments. To exclude patient specific variations caused by lymphopenia, we focused the analysis on the relative values.

The relative frequency of lymphocytes among leukocytes was significantly lower in critical versus moderate cases (Fig. S1H). We also observed a lower level of CD8^+^, higher CD4^+^ level and consequently a higher CD4^+^/CD8^+^ ratio in patients with severe and critical COVID-19 symptoms versus moderate (Fig. S1I-K), the difference was significant between moderate and severe patients.

### Decreased frequencies of lymphocytes with differentiated and activated cytotoxic phenotype is associated with critical COVID-19

Next, we performed a comparison of various T- and B-cell subsets between the different disease severities at the first visit (Fig. 2) and in follow-up (Fig. S3). We found lower frequencies of central memory CD4^+^ in patients with critical COVID-19 (Fig. 2A,B). Lower frequencies of terminally differentiated TEMRA CD4^+^ and CD8^+^ T-cells were found in patients with severe and critical compared to moderate COVID-19 (Fig. 2 C,D). Interestingly, for TEMRA CD8^+^ T-cells, the difference between the three cohorts was borderline significant (Kruskal-Wallis P=0.054); a pairwise examination of revealed a potential difference between critical and moderate patients (Mann-Whitney P=0.017). We further found lower frequencies of T-cells with an activated effector phenotype expressing HLA-DR and CD57 on CD4^+^ and CD8^+^ T-cells (Fig. 2E-H) in patients with critical disease compared to severe and moderate cases. While the differences for HLA-DR-expressing T-cells were statistically significant, the difference for CD8^+^CD57^+^ T cells showed only a tendency towards lower values with increasing COVID-19 severity (Kruskal-Wallis P=0.095). The difference was most marked between the moderate and the critical sub-cohorts (Mann-Whitney P=0.049). Interestingly, we found dramatically lower frequencies of CD11a-expressing CD4^+^ and CD8^+^ T-cells and a significant reduction of CD28^+^CD4^+^ T-cells in critical compared to severe and moderate COVID-19 (Fig. 2I-L). This alteration was obviously specific for the SARS-CoV-2 infection, as it was not observed in patients with pneumonia and sepsis on mechanical ventilation (data not shown).

In the B-cell compartment, we found a strong gradual reduction in the frequencies of transitional and marginal zone CD19^+^ cells in the patients with severe or critical symptoms (Fig. 2M-N), but not in class switched CD19^+^IgD^−^ plasmablasts (data not shown).

**Fig. 2:**
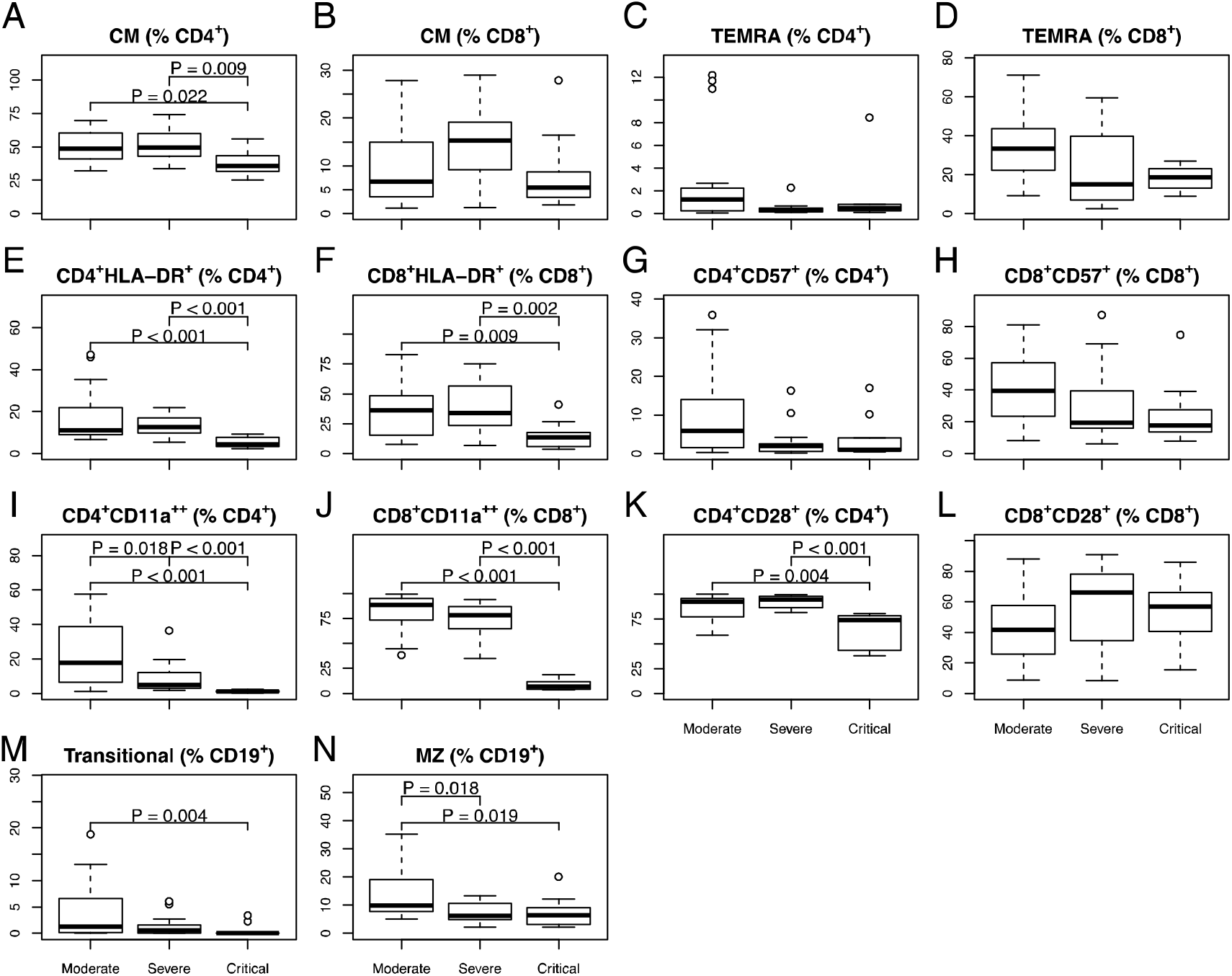
Decrease of lymphocyte frequencies with differentiated and activated cytotoxic phenotype in patients with severe or critical COVID-19. Peripheral blood from 53 patients with moderate, severe or critical COVID-19 manifestations was subjected to evaluation for differentiation and activation state of T- and B-cell subsets at the first visit using multiparametric flow cytometry. The subsets of the CD3^+^ T-cells and the CD19^+^ B lymphocyte were identified according to the gating strategy in Fig. S7-S9. (A-B) Frequency of central memory, defined as CCR7^+^CD45RA^−^, among CD4^+^ (A) and CD8^+^ (B) CD3^+^ T-cells. (C-D) Frequency of terminally differentiated effector T-cells (TEMRA), defined as CCR7^−^CD45RA^+^, among CD4^+^ (C) and CD8^+^ (D) CD3^+^ T-cells. (E-F) The frequency of HLA-DR expressing cells among CD4^+^ (E) and CD8^+^ (F) CD3^+^ T-cells. (G-H) The frequency of CD57 positive cells among CD4^+^ (G) and CD8^+^ (H) CD3^+^ T-cells. (I-J) The frequency of CD11a expressing cells among CD4^+^ (I) and CD8^+^ (J) CD3^+^ T-cells. (K-L) The frequency of CD28 expressing cells among CD4^+^ (K) and CD8^+^ (L) CD3^+^ T-cells. (M) The frequency of transitional B-cells defined as CD27^_^CD38^high^CD24^high^, among all CD19^+^IgM^+^IgD^+^ B-cells, and (N) the frequency of marginal zone (MZ) B-cells, defined as IgD^+^CD27^+^, among all CD19^+^ B-cells.

Taken together, the findings above demonstrate a loss of activated and differentiated effector T-cells in patients with severe and critical COVID-19. These differences remained stable albeit not always significant in follow up visit of moderate and severe COVID-19 patients (Fig. S3). The data on follow up analysis in critical COVID-19 patients with ARDS is of special importance. Six out of ten ARDS patients succumbed to the infection within 14 days after the study recruitment. At week three, however, all ARDS survivors displayed a recovery of the depleted CD11^++^ T-cell among CD4+ and CD8+ (Fig 3 A-B) including CD11a^++^ expressing HLA-DR, CD28, and CD57 molecules (Fig 3 C-F). In parallel, we also observed an increase of initially depleted TEMRA and HLA-DR^+^ CD8^+^ T-cells (Fig 3 G-H). At the time of writing, ARDS-survivors have been discharged from the ICU and no longer require mechanical ventilation.

**Fig. 3:**
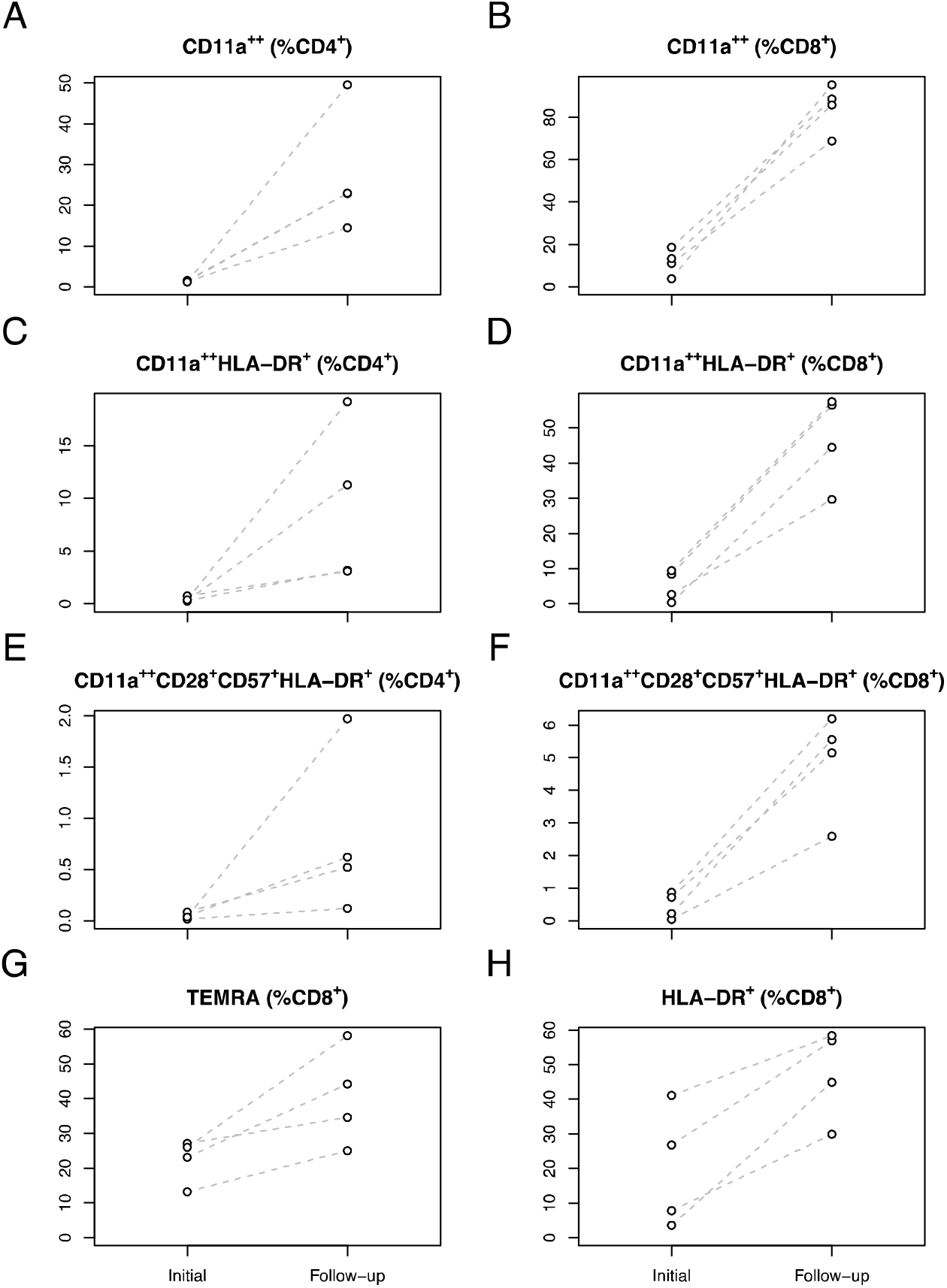
Recovery from ARDS is accompanied by the recovery of depleted T-cell subsets with activated differentiated effector phenotype. The kinetics of T-cell subsets with activated terminally differentiated effector phenotype was evaluated in four ARDS survivors. The subsets of the CD3 ^+^ T- cells were identified according to the gating strategy in Fig. S7 and S8. (A-B) The frequency of CD11a^++^ cells among CD4^+^ (A) and CD8^+^ (B) CD3^+^ T-cells. (C-D) The frequency of CD11a^++^HLA-DR^+^ expressing cells among CD4^+^ (C) and CD8^+^ (D) CD3^+^ T-cells. (E-F) The frequency of CD11a^++^HLA-DR^+^CD28^+^CD57^+^ expressing cells among CD4^+^ (E) and CD8^+^ (F) CD3^+^ T-cells. (G) Terminally differentiated effector T- cells (TEMRA), defined as CCR7^−^CD45RA^+^ among CD8^+^ T-cells. (H) Expression of HLA-DR among and CD8^+^ T-cells.

### Increased magnitude and functionality of SARS-CoV-2-reactive T-cells in patients with critical COVID-19

Given the observed loss of activated/differentiated T-cells in circulation of patients with critical symptoms, we wondered how this might influence the SARS-CoV-2 S-protein-specific T-cell immunity.

Among the patients with moderate symptoms, we found remarkably, albeit insignificantly, fewer patients with detectable SARS-CoV-2-reactive CD4^+^CD154^+^ T-cells compared to severe or critical cases (54.5% vs. 71.4% and 80.0%, respectively). The number of patients with detectable S-protein-reactive CD4^+^ T-cells increased in all groups to 76.9% (moderate), 100% (severe) and 81.8% (critical) after the follow-up visit (Table S4).

Interestingly, during the whole observation period, a lower percentage of patients with a moderate disease had detectable CD8^+^CD137^+^ T-cells (First visit: 54.5%, Follow-up: 38.5%), whereas the percentage of patients with a severe COVID-19 and detectable CD8^+^CD137^+^ T-cells increased from 42.9% at the first to 75% at the follow-up visit. In critically ill patients, the frequency was comparable to the level of the follow-up visit in severe diseases (80% and 81.8%).

Furthermore, we compared the magnitude of T-cell responses between the groups. We found a tendency towards higher frequency (Fig. 4A) and absolute counts (Fig. S4A) of S-protein reactive CD4^+^CD154^+^ in critical compared to moderate COVID-19. The magnitude of TNF-α, IFNγ and granzyme B producing CD4^+^CD154^+^ T-cells was slightly higher in the severe and critical versus moderate group (Fig. 4A), while for IL-2 producing CD4^+^CD154^+^ T cells, a significant difference was found. The magnitude of S-protein-reactive CD8^+^CD137^+^ T-cells was generally very low, although we found higher frequencies in critical COVID-19. We also detected a significantly higher frequency (Fig. 4B) and number (Fig. S4B) of IL-2 producing CD8^+^CD137^+^ T-cells, whereas the frequencies of TNF-α, IFNγ and granzyme B producing CD8^+^CD137^+^ T-cell were very low and comparable between the groups (Fig. 4B and Fig. S4B).

**Fig. 4:**
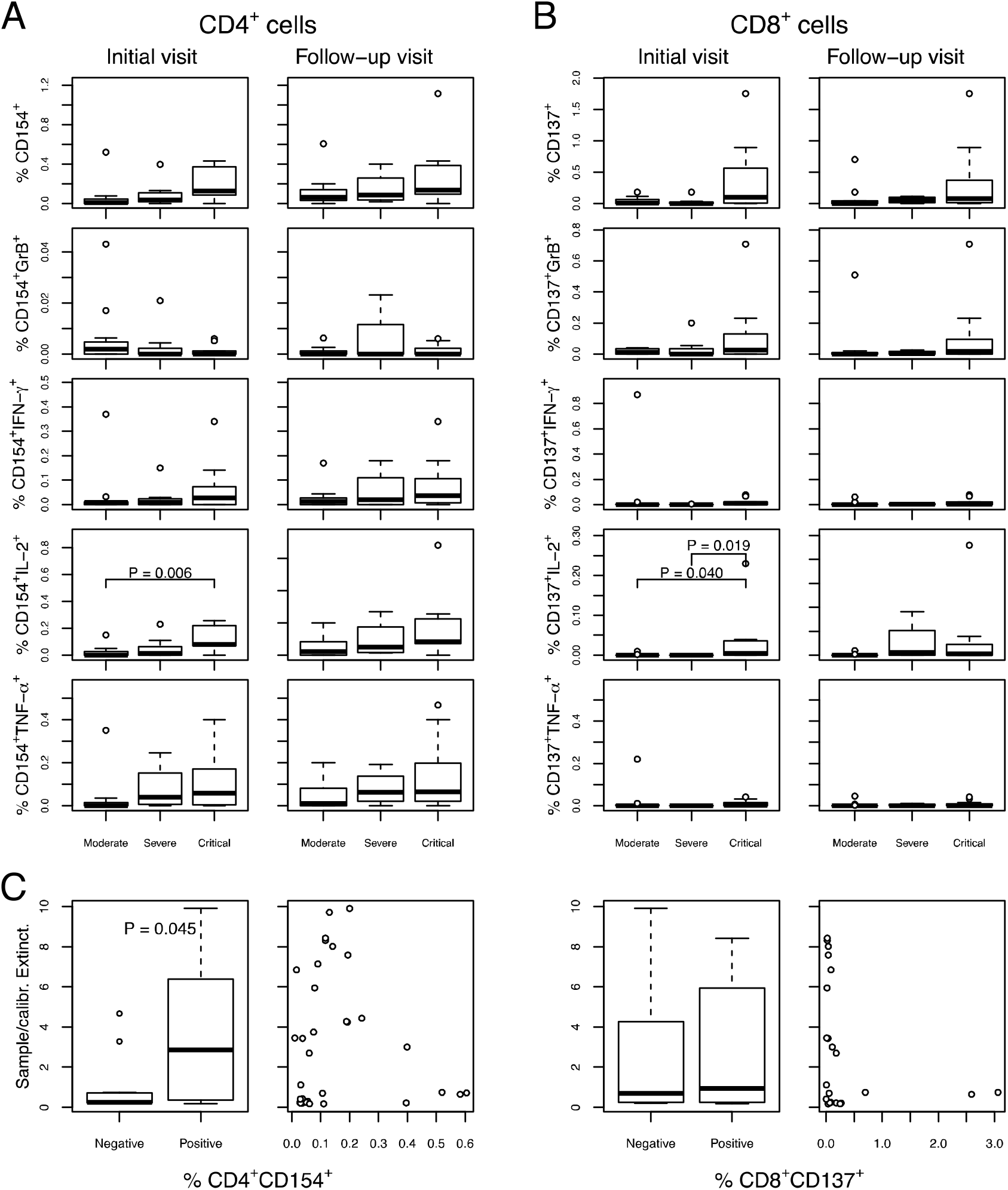
Increased magnitude of cytokine producing S-protein-reactive T-cells in patients with critical COVID-19. The presence and functional status of SARS-CoV-2-reactive T-cells was evaluated using PBMCs, isolated from the peripheral blood of patients with moderate (n=9), severe (n=8), or critical (n=10) COVID-19 manifestations. Defrosted PBMCs rested for 24 hours before treatment with overlapping peptide pools covering the SARS-CoV-2 S-protein. The cells were stimulated for a total of 16 hours and in the presence of Brefeldin A for the last 14 hours. The complete gating strategy is presented in Fig. S5. For critical COVID-19 patients, only a single time point was available. To ensure the comparability of the data with respect to the longer disease duration in critical COVID-19 patients, the comparison of the data in critically ill patients was performed also with the follow-up visit data in moderate and severe cases. (A) Frequency of CD4^+^CD154^+^ among CD4^+^ T-cells (first row) to the initial visit (left column) and follow-up visit (right column), and frequency of cells expressing granzyme B (GrB), INF-γ, IL-2, and TNF-α among CD4^+^CD154^+^ (row two to four). (B) Frequency of CD8^+^CD137^+^ among CD8^+^ T-cells (first row) to the initial visit (left column) and follow-up visit (right 13 column), and frequency of cells expressing granzyme B (GrB), INF-γ, IL-2, and TNF-α among CD8^+^CD137^+^ (row two to four). (C) Comparison of the relative titers of SARS-CoV-2 S-protein specific IgG antibodies, measured by ELISA and evaluated as ratio to an internal control for samples with detectable SARS-CoV-2 specific CD4^+^ T-cells (left) and SARS-CoV-2 specific CD8^+^ T-cells (right); correlation of the relative titers of SARS-CoV-2 with the counts of SARS-CoV-2 specific CD4^+^ T-cells (left) and SARS-CoV-2 specific CD8^+^ T-cells for samples with detectable T cell response.

Of interest, we found a correlation between humoral and cellular immunity: Samples with detectable SARS-CoV-2-reactive CD4^+^CD154^+^ T-cells had significantly higher antibody titers, independent of the COVID-19 severity and the sampling time point (Fig. 4C). Moreover, among the samples with detectable anti-SARS-CoV-2 CD4+ response the counts (Fig. S4C) of CD4^+^CD154^+^were significantly correlated with the magnitude of the humoral response. On the other hand, no significant effect was found for antiviral CD8^+^CD137^+^ T-cells.

## Discussion

Here, we present a comprehensive immune profiling of a cohort of 53 patients with different COVID-19 severity. Our data reveal an intriguing association between the quantitative composition and functionality of several immune cell subsets and the clinical manifestation of COVID-19 pointing to pathogenic immune reactivity in critical COVID-19. The most potent SARS-CoV-2 specific T cell immunity was detected in those patients with the worst lung injury. The temporary dramatic depletion of CD11a-expressing potent SARS-CoV-2 specific circulating T cells indicates that emigration from the vasculature with consecutive tissue invasion may constitute a central pathophysiological mechanism for tissue injury in COVID-19.

The impaired immune regulation and increased inflammation have been recently reported for patients with SARS-CoV-2-related severe respiratory failure (SRF) (5,13). Patients with SRF showed an IL-6-driven hyperinflammation and a T-, and B-cell lymphopenia (5). In agreement with these data, we show lower numbers of circulating T-, and B-cell subsets in patients with severe and critical as compared to moderate COVID-19. More importantly, critically ill patients had the lowest frequencies of T-cell subsets with advanced differentiation, activation and functional properties. The affected immune cell subsets are described to be involved in immune activation and cytotoxic response towards foreign antigens (14-16). The reason for the depletion of activated differentiated effector T-cells in circulation of patients with critical COVID-19 is not clear so far and can be explained either by activation-induced apoptosis or by inflammation-triggered cell migration. In this respect, the data on the dramatic depletion of CD11a-expressing CD4^+^ and CD8^+^ T-cells in patients with critical disease are relevant. CD11a is a key T-cell integrin, essential for T-cell activation and migration (17). In addition, CD57^+^T-cells are known as terminally differentiated, functionally competent memory/effector T lymphocytes, with high migratory capacities (15, 16). Notably, four out often ARDS patients recovered in follow up. This improvement in the clinical course of ARDS patients was accompanied by the increase of the depleted T-cell subsets. This points out that an inflammation-triggered lymphocyte migration (18,19) rather than apoptosis is responsible for the observed loss of activated terminally differentiated T-cell subsets. Moreover, previous data demonstrate that *in vivo* preactivated terminally differentiated TEMRA can migrate unspecifically to any inflammatory site providing cytotoxic effects (20). Supporting our hypothesis on inflammation-driven T-cell migration, the very recent review (21) reports on SARS-CoV-2-induced activation of IL-6 amplifiers. Leading to IL-6 and other cytokine release, it recruits activated T-cells and macrophages in the lesion (21). In line with this and other papers (5), we observed significantly higher levels of IL-6 in patients with severe and critical diseases (data not shown).

Thus, these findings provide substantial insight into the immunopathogenesis of COVID-19: In the observed patients hyperreactive and not insufficient cellular immunity is responsible for COVID-19 progression. We demonstrate an increased magnitude and functionality of SARS-CoV2 S-protein-reactive CD4^+^ and CD8^+^ T-cells in patients with critical and severe cases. The frequencies of SARS-CoV-2-reactive T-cells in critical COVID-19 are comparable or higher than the frequencies of other virus- or vaccine-reactive T-cells analyzed in our previous studies (7, 8,10, 22, 23). The magnitude of the S-protein-reactive T-cell response is also comparable with the first data on S-protein-reactive T-cells in patients with SARS-CoV-2-associated SRF (13). The reason for the higher number of SARS-CoV-2-reactive T-cells in critical cases has to be explored. It might be explained by a disturbed migration of the antigen-specific cells to the infected tissue leading to the impaired viral clearance, increased inflammation within infected tissue and unspecific migration of effector T-cells in this area through bystander activation. However, it is also possible that the composition of the peripheral immune cells mirrors the situation in the infected tissue, where the large number of antigen-specific effector T-cells leads to injury of the affected organ.

Independent of the reason for the higher magnitude of SARS-CoV2-reactive immunity, our data provide important clinical implications: Patients with severe and critical course mount a strong antiviral response. Although the protective capacity of SARS-CoV-2-reactive T-cells has to be evaluated, COVID-19 disease progression is obviously associated with a higher magnitude of inflammatory cytokine-producing cells. This finding delivers an immunological rationale for the use of immunosuppressive approaches at this stage of the disease. Concordantly, recent studies described a positive effect of anti-IL6 or anti-IL1 therapy in SRF patients (5, 24).

In conclusion, the data presented here are supportive of an immune pathogenesis as an underlying cause of COVID-19 severity. Additionally, the identified CD11a-based immune signature can represent a possible prognostic marker for disease progression. Since the proposed marker analysis is already offered by most immunodiagnostics laboratories, a multi-center evaluation is foreseeable and the marker can readily be utilized for monitoring in the current pandemic.

### Author contributions

Conceptualization: M.A., U.S. and N.B.; Data curation: K.P.; Formal analysis: M.A., A.B., C.J.T., T.R. and U.S.; Funding acquisition: T.H.W., O.W. and N.B.; Investigation: S.S., E.K., J.K., J.Z., P.W., S.K. and S.B.; Methodology: M.A., K.P., C.J.T. and T.R.; Project administration: U.S. and N.B.; Resources: K.P., A.D., F.S.S., B.H., M.J.K., T.B., C.T., C.W., S.D., U.D., T.H.W. and O.W.; Supervision: T.R., U.S. and N.B.; Visualization: A.B.; Writing – original draft: M.A., K.P., A.B., C.J.T., T.R., C.W., S.D., U.D., T.H.W., O.W., U.S. and N.B. The order of co-first authors is based on time spent on the project with alphabetical order of surnames resolving ties (M.A. and K.P.).

## Data Availability

Data available on request from the authors

## Acknowledgments

We want to express our deepest gratitude to the patients who donated their blood samples and clinical data for this project.

